# A prognostic signature based on ectopic reactivation of 8 tissue-specific genes in Diffuse Large B Cell Lymphoma

**DOI:** 10.64898/2026.04.23.26351580

**Authors:** Emilie Montaut, Vinciane Rainville, Patricia Betton-Fraisse, William Merre, Syrine Khedimallah, Jérôme Govin, Sophie Rousseaux, Saadi Khochbin, Fabrice Jardin, Phillipe Ruminy, Ekaterina Bourova-Flin, Anouk Emadali, Sylvain Carras

**Affiliations:** Univ. Grenoble Alpes, INSERM U1209, CNRS UMR 5309, Institute for Advanced Biosciences (IAB), Grenoble, France; Univ. Grenoble-Alpes, CHU Grenoble Alpes, Health Research and Innovation, Grenoble, France; INSERM U1245, Centre Henri Becquerel, University of Rouen, Rouen, France; Univ. Grenoble-Alpes, CHU Grenoble Alpes, Department of Molecular Biology, Grenoble, France; Department of Hematology, Centre Henri Becquerel, Rouen, France; Univ. Grenoble-Alpes, CHU Grenoble Alpes, Department Hematology, Grenoble, France

## Abstract

Diffuse Large B-cell lymphoma (DLBCL) is the most common aggressive lymphoma in the Western world. First-line immunochemotherapy fails in approximately 30–40% of patients, with refractory and relapse patients presenting a dismal prognosis. Currently, these high-risk patients cannot be accurately identified at diagnosis. Using statistical modeling and machine learning approaches applied to large public DLBCL datasets, we identified a novel predictive signature based on the reactivation of eight normally silent tissue-dependent genes associated with survival. We then developed a multiplex RT-MLPseq based assay, compatible with formalin-fixed paraffin-embedded (FFPE) samples and transferable into routine clinical practice, enabling analysis of expression of these eight genes and validated their prognosis impact in an independent real-life cohort. This signature could be integrated with current prognostic indices and molecular classifications to improve patient stratification and guide treatment selection toward a personalized theragnostic approach, thereby enhancing management of non-responder patients.

**Data Sharing Statement:** For access to original data, please contact

**Key points:**

- Ectopic activation of 8 tissue-specific genes defines a robust prognostic signature for survival stratification in DLBCL patients
- A FFPE-compatible RT-MLPseq assay enables clinical use and improves risk stratification beyond IPI and COO, especially in high-risk patients

## Introduction

Diffuse Large B-Cell Lymphoma (DLBCL) represents a heterogeneous group of lymphoid malignancies, accounting for approximately 30% of non-Hodgkin lymphoma^1^. Although the standard treatment for DLBCL, the R-CHOP regimen achieves long-term remission in 60-70% of patients, the remaining 30-40% present primary refractory disease or early relapse ^1^. Currently, risk is assessed using the IPI score (International Prognostic Index) ^2^. In 2002, Rosenwald *et al*. introduced a cell-of-origin (COO) classification of DLBCL based on gene expression profiling, distinguishing germinal center B-cell–like (GCB) from activated B-cell–like (ABC) subtypes ^3^. Recently, novel classifications based on mutational profiles were developed improving DLBCL molecular classification ^4–6^. However, these molecular classifiers have presently no value as predictive biomarkers of treatment response at the individual patient level. With the recent therapeutic advancements, such as CAR-T cells and bispecific anti-CD20 antibodies, the need for early identification of high-risk patients to guide risk adapted and innovative strategies is even more urgent.

Various epigenetic dysregulations ^7–10^ can lead to aberrant gene expression including reactivation of normally silent tissue-specific genes ^11^. These ectopic gene reactivations can be positively selected during oncogenic transformation when they support the acquired capabilities of tumor cells ^12,13^ and represent promising biomarkers for diagnosis, risk assessment or new therapeutic targets ^14^. To identify these events in DLBCL, we have adapted an innovative method based on *in silico* analysis of large series of public patient data using statistical modeling and machine learning ^15–17^. This approach presents the advantage of being robust by using several independent cohorts to identify and validate the most discriminative combination of genes associated with survival and enabled the identification of a robust signature, detectable with a clinically compatible assay, which could improve prognosis stratification in DLBCL patients.

## Methods

### Machine-Learning Discovery of Reactivated Tissue-Specific Genes Predicting Survival in DLBCL

We used normal samples gene expression profiles compiled from GTEx, NCBI SRA, and TCGA datasets to define tissue-specific genes as predominantly expressed in one tissue using a Z-score outlier detection method ^16^. After excluding genes mainly expressed in lymphoid tissues, those expressed at least five-fold higher in their predominant tissue than in lymphoid tissues were retained. Their abnormal expression in DLBCL was explored using the NCICCR-DLBCL cohort (n=481), defining reactivation when tumor expression exceeded the mean plus two standard deviations of normal lymphoid levels. A machine-learning framework, Ectopy ^16^, was then applied to determine optimal gene-expression thresholds associated with overall survival across training, validation and test cohorts (**Table S1**). Training in the EGAS00001002606 dataset (n=586) identified genes with survival-associated expression thresholds. Validation cohorts enabled the derivation of a gene classifier based aberrantly activated gene combinations showing consistent prognostic impact, whose performance was then assessed in independent test cohorts using log-rank and Cox proportional hazards analyses. Additional methodological details are given in the Supplementary Methods section.

### Development and validation of an RT-MLPseq-based LymphoGEC detection assay

RT-MLPseq assay was developed as previously described ^18,19^ using additional probes targeting LymphoGEC genes mRNA (**Table S2**). Detailed experimental procedures and data analysis methods are described in the Supplementary Methods section. To enable binary ON/OFF classification of genes, thresholds for gene activation were defined based on gene-specific percentile-rank thresholds established in the training cohort and applied to RTMLP-seq expression data obtained from 117 samples from a local cohort. External independent validation of the LymphoGEC signature was then performed using mRNA from FFPE tumoral biopsies (n=176) and the associated clinical data from R-CHOP-treated DLBCL patients enrolled in the RT3 trial (NCT03104478, Lymphoma Study Association) ^20^ (**Table S3)**.

## Results and discussion

To identify ectopic gene expression associated with prognosis in R-CHOP–treated DLBCL, we applied the Ectopy framework ^15–17^ (**Fig. 1A**). From a normal tissue expression metanalysis, 2,055 genes with exclusive or predominant expression in non-lymphoid tissues were identified. In the NCICCR-DLBCL cohort (481 cases), 617 of these genes were aberrantly reactivated (expression > mean + 2 SD) in ≥10% of DLBCL cases, confirming widespread ectopic expression of tissue-restricted genes in this disease, as previously reported in other neoplasms ^12,15–17,21^.

**Figure 1.**
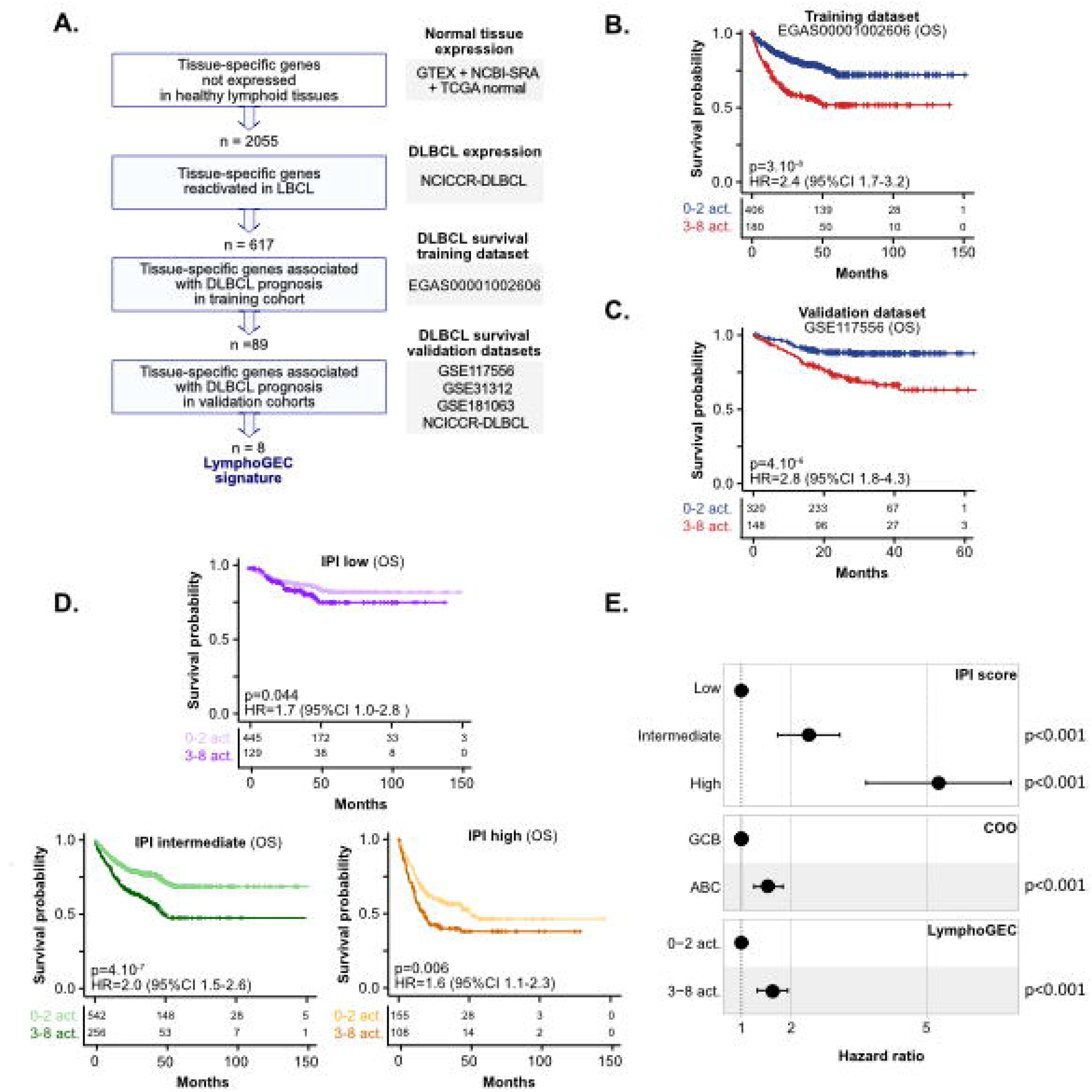
LymphoGEC predicts LBCL survival and refines IPI-based stratification. A. Framework for identification of ectopic gene reactivation associated with prognosis in DLBCL. B-C. Kaplan-Meier overall survival (OS) curves comparing patients presenting 0-2 and 3-8 LymphoGEC gene activation in training (EGA00001002606, B) and validation (GSE117556, C) datasets. D. Kaplan-Meier overall survival (OS) curves comparing patients presenting 0-2 and 3-8 LymphoGEC gene activation for low, intermediate and high IPI score LBCL cases (pooled datasets). Cox model p-values and hazard ratios (HR) including 95% confidence interval (95%CI) are reported. E. Forest plot of multivariate Cox proportional hazards regression overall survival analyses integrating IPI score, COO classification and LymphoGEC score (pooled datasets).

Using the EGAS00001002606 cohort (586 cases) as an independent training set, 89 genes whose activation status significantly stratified patient overall survival were identified. Percentile-based cutoffs were then applied to 4 independent validation cohorts (GSE117556, GSE31312, GSE181063 and NCICCR-DLBCL). Of these 89 genes, 8 genes reproducibly associated with prognosis in at least 2 validation cohorts were retained and combined into the LymphoGEC classifier. Although these genes display distinct tissue specificity and biological functions (**Fig. S1A, Table S4**), their cumulative activation stratified patients according to the number of reactivated genes (**Fig. S1B**), providing a robust prognostic signal across multiple independent cohorts and platforms.

Systematic testing of all possible combinations identified a threshold of ≥3 activated genes as providing the most significant prognostic impact in overall survival in the training dataset (p=3.10^-8^, **Fig. 1B**). This result was then confirmed in validation (**Fig 1C** and **Fig S1**), as well as in the test cohorts (**Fig. S1D**). We observed that LymphoGEC significantly stratified a higher-risk population in patients treated with R-CHOP as well as in those receiving bortezomib–R-CHOP (GSE117556, RB-CHOP arm), but not in patients treated with CHOP (GSE10846, CHOP arm). We next assessed whether the LymphoGEC prognostic signature presented an added value beyond IPI classification. Remarkably, LymphoGEC classification improved survival stratification in the intermediate (p=4.10^-7^) and high-risk categories (p=0.006, **Fig. 1C**), where clinical decision-making remains challenging. Multivariate Cox regression analyses adjusted for IPI and COO confirmed that LymphoGEC remained independently associated with overall survival (p<0.001, **Fig. 1D**), indicating that this molecular signature improves risk stratification beyond current prognostic models and may help guide risk adapted strategies in the era of novel agents and immunotherapies.

Next, we investigated DLBCL biological features based on LymphoGEC expression. We found that patients with more than 3 LymphoGEC activations were significantly enriched for the ABC subtype in 4 of 5 tested datasets (**Fig. S2A**), highlighting a preferential association with this molecular phenotype, despite their independence as prognosis factors. Moreover, expression correlation analysis between the 8 LymphoGEC genes revealed no evidence of coordinated regulation of the expression of these genes (**Fig. S2B**), indicating that their reactivation occurs independently rather than through a shared transcriptional program.

To assess the functional impact of LymphoGEC, we compared gene expression in GEC^+^ (≥4 reactivations) versus GEC^-^ (0 reactivations) DLBCL tumors across the two largest RNAseq cohorts (EGAS0000100606 and NCICCR-DLBCL and performed Gene Set Enrichment Analysis (GSEA). GEC^+^ tumors showed consistent enrichment of cell cycle, proliferation, and energy metabolism pathways, including MYC, E2F, and mTORC1 signaling, as well as fatty acid oxidation (**Fig. S3A**, up). Mitotic and protein metabolism programs, including the unfolded protein response and mTORC1-driven translation, were also upregulated in GEC+ DLBCL. The highly significant enrichment of dark-zone (DZsig) genes (NES=3, FWER<10^4^, **Fig. S3B**), recently defined as markers of high-risk DLBCL ^22^ also suggests the association of LymphoGEC activation with a hyperproliferative phenotype reminiscent of germinal center dark-zone B cells. GO analysis of commonly upregulated genes confirmed altered metabolism and cell homeostasis (**Fig. S3C**). In contrast, GEC-tumors were notably enriched for immune-related signatures (**Fig. S3A**, down), morphogenesis, and collective cell movement programs (**Fig. S3D**), supporting a model in which GEC^+^ cases exhibit loss of cell identity, deregulated differentiation, and immune evasion. Collectively, these data suggest that LymphoGEC captures a global biological shift toward anabolic metabolism and cell-cycle acceleration, two classical hallmarks of treatment-resistant disease ^23^, potentially underlying its dismal prognostic impact.

To enable routine-compatible detection of LymphoGEC gene reactivation, we developed a multiplex RT-MLPseq assay compatible with FFPE samples incorporating probes specific of the eight LymphoGEC genes. Cut-off values were defined based on gene-specific percentile-rank thresholds established in the training cohort and applied to RTMLP-seq expression data obtained from 117 DLBCL samples.

LymphoGEC gene reactivation was then analyzed in the RT3 prospective cohort (**Table S2)** by applying these RT-MLPseq counts thresholds. Using this alternative gene expression detection method in an independent cohort, we confirmed a cumulative association between the number of LymphoGEC gene activations and adverse prognosis (**Fig. S4A**). Applying the threshold of at least three activated genes previously identified resulted in a highly significant separation of both progression-free (p=0.001, **Fig. 2A)** and overall survival groups (p=0.03, **Fig S4B**) and markedly improved stratification within the intermediate-risk IPI category (**Fig. 2B**). We also confirmed enrichment of ABC DLBCL in GEC+ patients (**Fig. S4C**), lack of correlation among LymphoGEC genes (**Fig. S4D**), and its independence from IPI as a prognostic factor (**Fig. 2C**), further validating LymphoGEC relevance and transferability.

**Figure 2.**
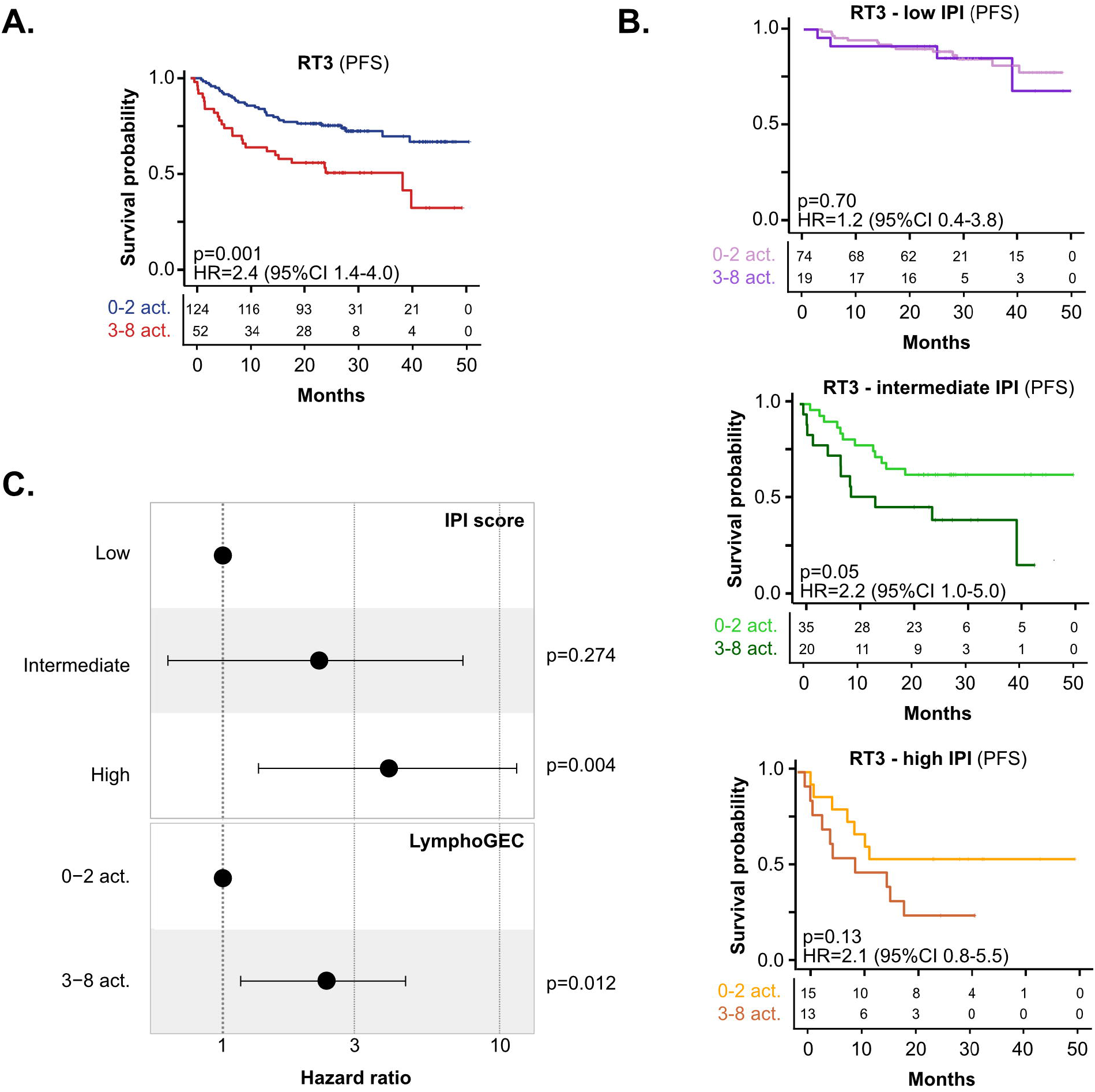
LymphoGEC detection by RT-MLPseq predicts LBCL survival and refines IPI-based stratification in an independent LBCL real-life cohort. **A**. Kaplan-Meier progression free survival (PFS) curves comparing patients presenting 0-2 and 3-8 LymphoGEC gene activation in the RT3 cohort. **B**. Kaplan-Meier progression free survival (PFS) curves comparing patients presenting 0-2 and 3-8 LymphoGEC gene activation in the RT3 cohort for low, intermediate and high IPI score LBCL cases. Cox model p-values and hazard ratios (HR) including 95% confidence interval (95%CI) are reported. **C**. Forest plot of multivariate Cox proportional hazards regression analyses integrating the IPI and LymphoGEC scores.

Further investigation is needed to clarify the mechanisms underlying LymphoGEC gene reactivation and to better define the functional role of these genes in lymphoma biology, both of which will be important for fully assessing their therapeutic potential.

Yet, LymphoGEC captures a biologically relevant transcriptional program through an original approach and highlights a new facet of LBCL biology. It also emerges as a robust classifier that independently predicts survival in LBCL. Its implementation through a multiplex RT-MLPseq assay enables rapid and scalable detection in routine samples, supporting integration into diagnostic workflows and its potential to inform risk-adapted and precision therapeutic strategies.

## Data Availability

All data produced in the present study are available from the corresponding author upon request.

## Acknowledgements

We gratefully acknowledge Dr. Sandrine Roulland for her careful review of the manuscript and her insightful comments. This work was supported by internal funding from Grenoble University Hospital (Appel d’Offre Interne Recherche et Innovation – EPIC project) and by the Pôle Universitaire d’Innovation (PUI) Grenoble Alpes (pre-maturation program C7H-PUI24A10-LYMPHOGEC). We also thank LYSA/LYSARC for their support in the management of RT3 samples and associated clinical data, and the DRCI of CHU Grenoble Alpes for their valuable contribution to the project.

## Authorship Contributions

EM set up all bioinformatic and statistical analyses pipelines; EM, PBF, WM and SKhe performed bioinformatic analyses and designed figures; VR and PR designed and performed RT-MLPseq experiments; PR and FJ developed RT-MLPseq assay and provided access to validation samples; SR, JG, SKho, EBF, AE and SC conceived the study; AE and SC coordinated the study; EM, AE and SC: wrote the manuscript. All authors reviewed and edited the manuscript.

## Bibliography

1. Sehn LH, Salles G. Diffuse Large B-Cell Lymphoma. N Engl J Med. 2021;384(9):842–858. doi:10.1056/NEJMra2027612

2. Sehn LH, Berry B, Chhanabhai M, et al. The revised International Prognostic Index (R-IPI) is a better predictor of outcome than the standard IPI for patients with diffuse large B-cell lymphoma treated with R-CHOP. Blood. 2007;109(5):1857–1861. doi:10.1182/blood-2006-08-038257

3. Rosenwald Andreas, Wright George, Chan Wing C., et al. The Use of Molecular Profiling to Predict Survival after Chemotherapy for Diffuse Large-B-Cell Lymphoma. N Engl J Med. 2002;346(25):1937–1947. doi:10.1056/NEJMoa012914

4. Chapuy B, Stewart C, Dunford AJ, et al. Molecular Subtypes of Diffuse Large B-cell Lymphoma are Associated with Distinct Pathogenic Mechanisms and Outcomes. Nat Med. 2018;24(5):679–690. doi:10.1038/s41591-018-0016-8

5. Schmitz Roland, Wright George W., Huang Da Wei, et al. Genetics and Pathogenesis of Diffuse Large B-Cell Lymphoma. N Engl J Med. 2018;378(15):1396–1407. doi:10.1056/NEJMoa1801445

6. Wright GW, Huang DW, Phelan JD, et al. A Probabilistic Classification Tool for Genetic Subtypes of Diffuse Large B Cell Lymphoma with Therapeutic Implications. Cancer Cell. 2020;37(4):551–568.e14. doi:10.1016/j.ccell.2020.03.015

7. Diacofotaki A, Loriot A, De Smet C. Identification of Tissue-Specific Gene Clusters Induced by DNA Demethylation in Lung Adenocarcinoma: More Than Germline Genes. Cancers. 2022;14(4):1007. doi:10.3390/cancers14041007

8. Jeong HM, Kwon MJ, Shin YK. Overexpression of Cancer-Associated Genes via Epigenetic Derepression Mechanisms in Gynecologic Cancer. Front Oncol. 2014;4:12. doi:10.3389/fonc.2014.00012

9. Rao M, Chinnasamy N, Hong JA, et al. Inhibition of Histone Lysine Methylation Enhances Expression of Cancer-Testis Genes in Lung Cancer Cells: Implications for Adoptive Immunotherapy of Cancer. Cancer Res. 2011;71(12):4192–4204. doi:10.1158/0008-5472.CAN-10-2442

10. Glancy E, Choy N, Eckersley-Maslin MA. Bivalent chromatin: a developmental balancing act tipped in cancer. Biochem Soc Trans. 2024;52(1):217–229. doi:10.1042/BST20230426

11. Axelsen JB, Lotem J, Sachs L, Domany E. Genes overexpressed in different human solid cancers exhibit different tissue-specific expression profiles. Proc Natl Acad Sci. 2007;104(32):13122–13127. doi:10.1073/pnas.0705824104

12. Rousseaux S, Wang J, Khochbin S. Cancer hallmarks sustained by ectopic activations of placenta/male germline genes. Cell Cycle Georget Tex. 2013;12(15):2331–2332. doi:10.4161/cc.25545

13. Wang J, Rousseaux S, Khochbin S. Sustaining cancer through addictive ectopic gene activation. Curr Opin Oncol. 2014;26(1):73–77. doi:10.1097/CCO.0000000000000032

14. Wang ML, Barrientos JC, Furman RR, et al. Zilovertamab Vedotin Targeting of ROR1 as Therapy for Lymphoid Cancers. NEJM Evid. 2022;1(1):EVIDoa2100001. doi:10.1056/EVIDoa2100001

15. Bourova-Flin E, Derakhshan S, Goudarzi A, et al. The combined detection of Amphiregulin, Cyclin A1 and DDX20/Gemin3 expression predicts aggressive forms of oral squamous cell carcinoma. Br J Cancer. 2021;125(8):1122–1134. doi:10.1038/s41416-021-01491-x

16. Jacquet E, Chuffart F, Vitte AL, et al. Aberrant activation of five embryonic stem cell-specific genes robustly predicts a high risk of relapse in breast cancers. BMC Genomics. 2023;24(1):1. doi:10.1186/s12864-023-09571-3

17. Peng LJ, Zhou YB, Geng M, et al. Ectopic expression of a combination of 5 genes detects high risk forms of T-cell acute lymphoblastic leukemia. BMC Genomics. 2022;23(1):1. doi:10.1186/s12864-022-08688-1

18. Bobée V, Drieux F, Marchand V, et al. Combining gene expression profiling and machine learning to diagnose B-cell non-Hodgkin lymphoma. Blood Cancer J. 2020;10(5):59. doi:10.1038/s41408-020-0322-5

19. Wang J, Yang X, Chen H, et al. A high-throughput method to detect RNA profiling by integration of RT-MLPA with next generation sequencing technology. Oncotarget. 2017;8(28):46071–46080. doi:10.18632/oncotarget.17551

20. Copie Bergman C, Bohers E, Dartigues-Cuillères P, et al. Real Time Pathological and Molecular Characterization of Aggressive B-Cell Lymphomas Based on a National Network. a Lysa Project. Blood. 2020;136:22–23. doi:10.1182/blood-2020-141953

21. Wang J, Mi JQ, Debernardi A, et al. A six gene expression signature defines aggressive subtypes and predicts outcome in childhood and adult acute lymphoblastic leukemia. Oncotarget. 2015;6(18):16527–16542. doi:10.18632/oncotarget.4113

22. Alduaij W, Collinge B, Ben-Neriah S, et al. Molecular determinants of clinical outcomes in a real-world diffuse large B-cell lymphoma population. Blood. 2023;141(20):2493–2507. doi:10.1182/blood.2022018248

23. Ward PS, Thompson CB. Metabolic Reprogramming: A Cancer Hallmark Even Warburg Did Not Anticipate. Cancer Cell. 2012;21(3):297–308. doi:10.1016/j.ccr.2012.02.014

