## Supplemental material for "A prognostic signature based on ectopic reactivation of 8 tissue-specific genes in Diffuse Large B Cell Lymphoma"

### Supplemental data

#### Supplemental methods

**Public gene expression databases.** DLBCL cancer datasets including gene expression and clinical data were obtained from TCGA/GDC portal (NCICCR-DLBCL), European Genome-phenome Archive (EGA) at the European Bioinformatics Institute (EBI) (EGAS00001002606, through a data access agreement) [1] and GEO repository (GSE117556, GSE31312, GSE181063, GSE10846 [CHOP and R-CHOP arms], GSE117556 [R-CHOP and RB(Rituximab+Bortezomib)-CHOP arms], GSE53786, GSE32918 and GSE87371). The detailed description of the datasets is presented in **Table S1**. For RNA-seq datasets, we used RPM-normalized counts (provided by the GDC Data Portal for NCICCR-DLBCL or calculated in-house from raw counts for EGAS00001002606) [2]. The RPM values were pseudo log-transformed (log2[1 + RPM]). Microarray datasets (Affymetrix Human Genome Arrays U133 Plus 2.0 or Illumina HumanHT-12 WG-DASL V4.0 R2) were normalized using Robust Multi-array Average (RMA) method [3] and log2-transformed.

**Identification of tissue-specific genes reactivated in DLBCL.** Gene expression profiles in normal tissues were obtained from a database combining GTEX, NCBI sequence Read Archive normal samples (datasets PRJNA280600, PRJEB4337, PRJEB2445, PRJNA270632, GSE70741, GSE53096), and TCGA non-tumor samples expression data (total sample size of the pooled dataset: 3686 samples, 45 tissues). Using the average expression levels in available tissue types, we identified tissue-specific genes as predominantly expressed in one tissue and not expressed, or lowly expressed, in all other somatic tissues. To detect the predominant tissues, for each gene, we applied an outlier detection technique based on the Z-score, as previously described [4]. After excluding genes with predominant expression in lymphoid tissues (lymph node, spleen and tonsil), we selected those whose expression in the predominant tissue was at least five-fold higher than in lymphoid tissue. Next, we calculated the frequency of aberrant expression of these tissue-specific genes in DLBCL using the NCICCR-DLBCL dataset, in which public RNA-seq data with sufficiently large sample size (n=481) were available. A gene was defined as reactivated when its expression in DLBCL exceeded the mean + 2 standard deviations of the expression level observed in normal lymphoid tissues. The proportion of tumor samples exceeding this threshold was calculated, and the 617 genes reactivated in more than 10% of DLBCL patients were retained for further analysis.

**Machine-learning identification of the optimal combination of reactivated tissue-specific genes for predicting DLBCL survival.** Survival analyses were then performed to explore the association between ectopic gene expression and overall survival based. A machine learning framework named Ectopy [4] was applied to determine the optimal activation threshold for each gene across DLBCL cohorts, that were divided into training, validation and test datasets. Ectopy aims at identifying robust prognostic biomarkers by learning gene-expression thresholds associated with survival in a large training cohort, validating their stability across heterogeneous cohorts, and combining consistent genes into a classifier that stratifies patients by survival. In the training step, Ectopy identifies genes whose expression levels can stratify patients into groups with significantly different survival by systematically testing expression thresholds in a large reference cohort and retaining only stable and statistically robust thresholds through cross-validation. In the validation step, these thresholds are transferred to independent and heterogeneous cohorts to select genes that show consistent prognostic value across multiple datasets. In the test step, the validated genes are combined into a classifier, which is then evaluated in fully independent cohorts to confirm its ability to predict patient prognosis. The Ectopy pipeline was applied to DLBCL data as described below.

*Training.* The EGAS00001002606 dataset was selected as training cohort due to its large sample size and high-quality RNA-seq data. For each gene, all possible thresholds (in the range from $15^{th}$ to 85th percentiles, with a step of a half of percentile) were analyzed using the log rank statistical test between the group of tumors in which the gene was aberrantly activated ("ON") and the group of tumors without aberrant gene reactivation ("OFF"), in full and cross-validated datasets. P-values were Benjamini–Hochberg–adjusted; significance required $p < 0.05$, $FDR < 0.2$, $HR > 1$. The most stable significant threshold across cross-validations was retained. In the training step, 89 candidate genes with at least one significantly survival-associated threshold were selected.

*Validation.* The thresholds obtained for the 89 selected genes were converted to percentile ranks (42% for OXTR; 38% for DNAJC6; 25% for TRIM9; 18% for EHHADH; 16.5% for MT1H; and 15% for OTX1, FOXA3, and PDCH9) and applied to four validation datasets NCICCR-DLBCL, GSE117556, GSE31312 and GSE181063. In this step, we used heterogeneous cohorts in terms of composition, sample size and technology to retain candidate biomarkers with the most consistent and stable prognostic value across multiple datasets. Log-rank tests were performed between the ON and OFF groups in four validation cohorts, using the thresholds calculated in the training step. Then, the 89 genes were ranked accordingly to the obtained p-values and hazard ratios in all validation cohorts. Candidate genes ($p < 0.05$ in at least two out of five cohorts) were then analyzed by groups to define the combination that most significantly discriminate against survival groups. This led to the identification of an 8-genes expression classifier, that we called LymphoGEC, which stratifies patients according to the number of aberrantly activated genes. Higher number of aberrant activations within the panel of these 8 genes is associated with a poorer patients' prognosis.
*Test.* Finally, the combined LymphoGEC tool was tested in several independent DLBCL cohorts, using the log rank test and Cox proportional hazard model. The test cohorts include GSE10846 (R-CHOP treatment arm), GSE10846 (CHOP treatment arm), GSE117556 (RB-CHOP treatment arm) and a merge of the GSE53786, GSE32918 and GSE87371 datasets which were pooled to increase sample size.

**Survival analysis.** Overall and progression-free survival were evaluated using both univariate and multivariate approaches. For the univariate analysis, survival curves were estimated by the Kaplan–Meier method, and differences between groups were assessed using Cox proportional hazards regression. To minimize the impact of late events, outcomes occurring beyond 5 years of follow-up were censored. For the multivariate analysis, Cox proportional hazards regression models were constructed to adjust for potential confounding variables and to identify independent prognostic factors. Hazard ratios (HR) with corresponding 95% confidence intervals are reported.

**Differential gene expression analysis**. Analyses were performed with R release 4.1.0. Differential genes expression analysis between patients with 0 LymphoGEC activation and patients with 4-8 gene activations was carried out using the DESeq2 package [5]. Differences between groups were assessed using the Wald test with Benjamini–Hochberg correction for multiple testing. Genes with an adjusted p-value $< 0.05$ and a log2 fold change $< -1$ or $> 1$ were considered significantly differentially expressed. Enrichment analyses were performed using GSEA software [6] and WebGestalt analysis toolkit [7] with default parameters**.**

**RT-MLPseq.** RT-MLPseq assay was performed on RNA samples, as previously described [8,9]. Briefly, total RNAs, extracted from formalin-fixed paraffin-embedded (FFPE) biopsies and converted into cDNA by reverse transcription using a M-MLV Reverse transcriptase and random hexamers to avoid 3' end bias (Invitrogen, Carlsbad, CA). cDNA are incubated 1 h at 60 °C with a mix of ligation dependent PCR oligonucleotides probes previously described [10] added with probes targeting LymphoGEC genes mRNA (**Table S2**), including universal adaptor sequences and random sequences of 7 nucleotides as unique molecular identifiers (UMI) in 1× SALSA MLPA buffer (MRC Holland, Amsterdam, the Netherlands), ligated using the thermostable SALSA DNA ligase (MRC Holland, Amsterdam, the Netherlands), and amplified by PCR using barcoded primers containing P5 and P7 adaptor sequences with the Q5 hotstart high fidelity master mix (NEB, Ipswich, MA). Amplification products are next purified using AMPure XP beads (Beckman Coulter, Brea, CA) and analyzed using a MiSeq sequencer (Illumina, San Diego, CA). Sequencing reads are de-multiplexed using the index sequences introduced during PCR amplification, aligned with the sequences of the probes and counted. All results are normalized according to the UMI sequences to avoid PCR amplification bias. Results are considered interpretable when at least 5000 different UMI (corresponding to the sum of all the UMI, for all the markers) are detected, allowing the evaluation of an average range of 1–40 for each marker.

**Supplemental references**


1. Reddy A, Zhang J, Davis NS, et al. Genetic and Functional Drivers of Diffuse Large B Cell Lymphoma. *Cell*. 2017;171(2):481-494.e15. doi:10.1016/j.cell.2017.09.027

2. Trapnell C, Williams BA, Pertea G, et al. Transcript assembly and quantification by RNA-Seq reveals unannotated transcripts and isoform switching during cell differentiation. *Nat Biotechnol*. 2010;28(5):511-515. doi:10.1038/nbt.1621

3. Irizarry RA, Hobbs B, Collin F, et al. Exploration, normalization, and summaries of high density oligonucleotide array probe level data. *Biostat Oxf Engl*. 2003;4(2):249-264. doi:10.1093/biostatistics/4.2.249

4. Jacquet E, Chuffart F, Vitte AL, et al. Aberrant activation of five embryonic stem cell-specific genes robustly predicts a high risk of relapse in breast cancers. *BMC Genomics*. 2023;24(1):1. doi:10.1186/s12864-023-09571-3

5. Love MI, Huber W, Anders S. Moderated estimation of fold change and dispersion for RNA-seq data with DESeq2. *Genome Biol*. 2014;15(12):12. doi:10.1186/s13059-014-0550-8

6. Subramanian A, Tamayo P, Mootha VK, et al. Gene set enrichment analysis: A knowledge-based approach for interpreting genome-wide expression profiles. *Proc Natl Acad Sci U S A*. 2005;102(43):15545-15550. doi:10.1073/pnas.0506580102

7. Elizarraras JM, Liao Y, Shi Z, Zhu Q, Pico AR, Zhang B. WebGestalt 2024: faster gene set analysis and new support for metabolomics and multi-omics. *Nucleic Acids Res*. 2024;52(W1):W415-W421. doi:10.1093/nar/gkae456

8. Bobée V, Drieux F, Marchand V, et al. Combining gene expression profiling and machine learning to diagnose B-cell non-Hodgkin lymphoma. *Blood Cancer J*. 2020;10(5):59. doi:10.1038/s41408-020-0322-5

9. Wang J, Yang X, Chen H, et al. A high-throughput method to detect RNA profiling by integration of RT-MLPA with next generation sequencing technology. *Oncotarget*. 2017;8(28):46071-46080. doi:10.18632/oncotarget.17551

10. Copie Bergman C, Bohers E, Dartigues-Cuillères P, et al. Real Time Pathological and Molecular Characterization of Aggressive B-Cell Lymphomas Based on a National Network. a Lysa Project. *Blood*. 2020;136:22-23. doi:10.1182/blood-2020-141953

#### Supplemental Table 1: DLBCL public data sets description

| Variables | Datasets | | | | | | | | | | |
|---|---|---|---|---|---|---|---|---|---|---|---|
| | **NCICCR-DLBCL** | **EGAS00001002606** | **GSE 117556 R-CHOP** | **GSE 31312** | **GSE 181063** | **GSE 10846 R-CHOP** | **GSE 10846 CHOP** | **GSE 117556 RB-CHOP** | **GSE 53786** | **GSE 32918** | **GSE 87371** |
| **Case number** | 481 * | 586 | 468 | 470 | 381** | 233 | 180 | 456 | 70 | 140 | 76 |
| **Number of events (death)** | 98 | 180 | 81 | 170 | 180 | 60 | 104 | 72 | 15 | 62 | 24 |
| **Date** | 2018 | 2018 | 2011 | 2011 | 2021 | 2005 | 2005 | 2011 | 2014 | 2011 | 2016 |
| **Technology** | RNA-Seq | RNA-Seq | Micro-array | Micro-array | Micro-array | Micro-array | Micro-array | Micro-array | Micro-array | Micro-array | Micro-array |
| **Treatment** | R-CHOP | R-CHOP | R-CHOP | R-CHOP | R-CHOP | R-CHOP | CHOP | RB-CHOP | R-CHOP | R-CHOP | R-CHOP |
| **Age (median)** | 61 | 62 | 66 | 63 | 66 | 61 | 65 | 63 | 61 | 69 | 63 |
| **IPI score (Low/Intermediate/High)** | 81/89/23 | 158/244/100 | 126/260/82 | 169/197/58 | 25/29/3 | 82/66/16 | NA | NA | 7/26/5 | NA | 18/34/24 |
| **ABC/GCB subtype** | NA | 243/255 | 129/276 | 199/227 | 15/25 | 93/107 | NA | NA | 30/30 | NA | 27/31 |

*234 with survival data, **378 with survival data, NA: non available

#### Supplemental Table 2: RT-MLPseq probes

| Probe name | Probe sequence |
|---|---|
| DNAJC6E4G | GTGCCAGCAAGATCCAATCTAGANNNNNNNCGAACTGCCAAGTTTCACAGCCGG |
| DNAJC6E5D | GTCTCAGAATGCAGTTGGCCCATTAGGCTCCAACCCTTAGGGAACCC |
| EHHADHE6G | GTGCCAGCAAGATCCAATCTAGANNNNNNNGGCCTGTCTCCTCAGTTGGTGTTGTTG |
| EHHADHE7D | GCTTGGGAACAATGGGCCGAGTCCAACCCTTAGGGAACCC |
| OXTRE3G | GTGCCAGCAAGATCCAATCTAGANNNNNNNTGCCAACGCGCCCAAGGAAG |
| OXTRE4D | CCTCGGCCTTCATCATCGTCATGCTTCCAACCCTTAGGGAACCC |
| OTX1E4G | GTGCCAGCAAGATCCAATCTAGANNNNNNNAAGATCAACCTGCCGGAGTCTAGAGTCCAG |
| OTX1E5D | GTCTGGTTCAAGAACCGCCGCGTCCAACCCTTAGGGAACCC |
| TRIM9E6G | GTGCCAGCAAGATCCAATCTAGANNNNNNNGGCAACGGTGGTCAATTCCGG |
| TRIM9E7D | GAGGTGTATGTGGGGAAGGAGACAATGTGTCCAACCCTTAGGGAACCC |
| MT1HE1G | GTGCCAGCAAGATCCAATCTAGANNNNNNNGGACCCCAACTGCTCCTGCGA |
| MT1HE1D | GGCTGGTGGCTCCTGCGCCTTCCAACCCTTAGGGAACCC |
| PCDH9E4G | GTGCCAGCAAGATCCAATCTAGANNNNNNNAGCAGATGGCAACTCTGATCCCAACTCTG |
| PCDH9E5D | ATGGGCCTTTGGGTCCCCGAGTCCAACCCTTAGGGAACCC |
| FOXA3E1G | GTGCCAGCAAGATCCAATCTAGANNNNNNNCTACTACCCGGAGGCGGGCGAG |
| FOXA3E2D | GTCTACTCGCCGGTGACCCCAGTGTCCAACCCTTAGGGAACCC |

#### Supplemental Table 3: RT3 cohort description

| | Total (n=176)[1] | GEC 0-2 (n=124)[1] | GEC 3-8 (n=52)[1] | Comparison test (p-value)[2] |
|---|---|---|---|---|
| **Follow-up time (months)** | 24 (14, 29) | 25 (20, 30) | 22 (6, 26) | **0.004** |
| **Age at diagnosis** | 65 (53, 73) | 61 (50, 71) | 70 (60, 78) | **<0.001** |
| **Sex** | | | | 0.5 |
| Female | 68 (39%) | 50 (40%) | 18 (35%) | |
| Male | 108 (61%) | 74 (60%) | 34 (65%) | |
| **IPI stage** | | | | 0.2 |
| 0-1 (Low risk) | 51 (29%) | 40 (32%) | 11 (21%) | |
| 2-3 (Intermediate risk) | 82 (47%) | 57 (46%) | 25 (48%) | |
| 4-5 (High risk) | 43 (24%) | 27 (22%) | 16 (31%) | |
| **COO status** | | | | **<0.001** |
| ABC | 90 (52%) | 51 (41%) | 39 (76%) | |
| GCB | 83 (48%) | 71 (58%) | 12 (24%) | |
| unclassified | 1 (0.6%) | 1 (0.8%) | 0 (0%) | |
| **MYC - BCL2** | | | | 0.3 |
| MYC_R & BCL2_R | 7 (17%) | 4 (12%) | 3 (38%) | |
| MYC_R - BCL2_R | 33 (80%) | 28 (85%) | 5 (63%) | |
| Not researched | 1 (2.4%) | 1 (3.0%) | 0 (0%) | |

[1]Median (Q1, Q3); n (%)

[2]Wilcoxon rank sum test; Pearson's Chi-squared test; Fisher's exact test

#### Supplemental Table 4: Tissue specificity, activity and function of the 8 LymphoGEC genes

| Gene name | Main tissue expression | Biological Activity | Cellular Function |
|---|---|---|---|
| **DNAJC6** | Brain | Tyrosine phosphatase activity, promotes uncoating of clathrin-coated vesicle | Clathrin-mediated endocytosis |
| **EHHADH** | Liver | Enoyl-CoA Hydratase and 3-Hydroxyacyl-CoA Dehydrogenase | Lipid metabolism, peroxisomal beta-oxidation pathway |
| **OXTR** | Breast | G-protein coupled receptor | Phosphatidylinositol-calcium signaling (parturition and lactation) |
| **OTX1** | Skin | Homeodomain-containing transcription factor | Brain and sense organ development |
| **TRIM9** | Brain | RING-type E3 ubiquitin-protein ligase, proteasome degradation | Regulation of neuronal functions, negative regulation of NF-kappaB activity |
| **MT1H** | Peritoneum | Heavy metals binding, transcriptionally regulated by glucocorticoids | Cellular response to zinc, cadmium and copper |
| **PCDH9** | Liver | Transmembrane proteins containing cadherin domains | Cell adhesion in neural tissue, inhibits EMT and cell adhesion |
| **FOXA3** | Brain | Pioneer transcription factor | Transcriptional activators for liver-specific transcripts |

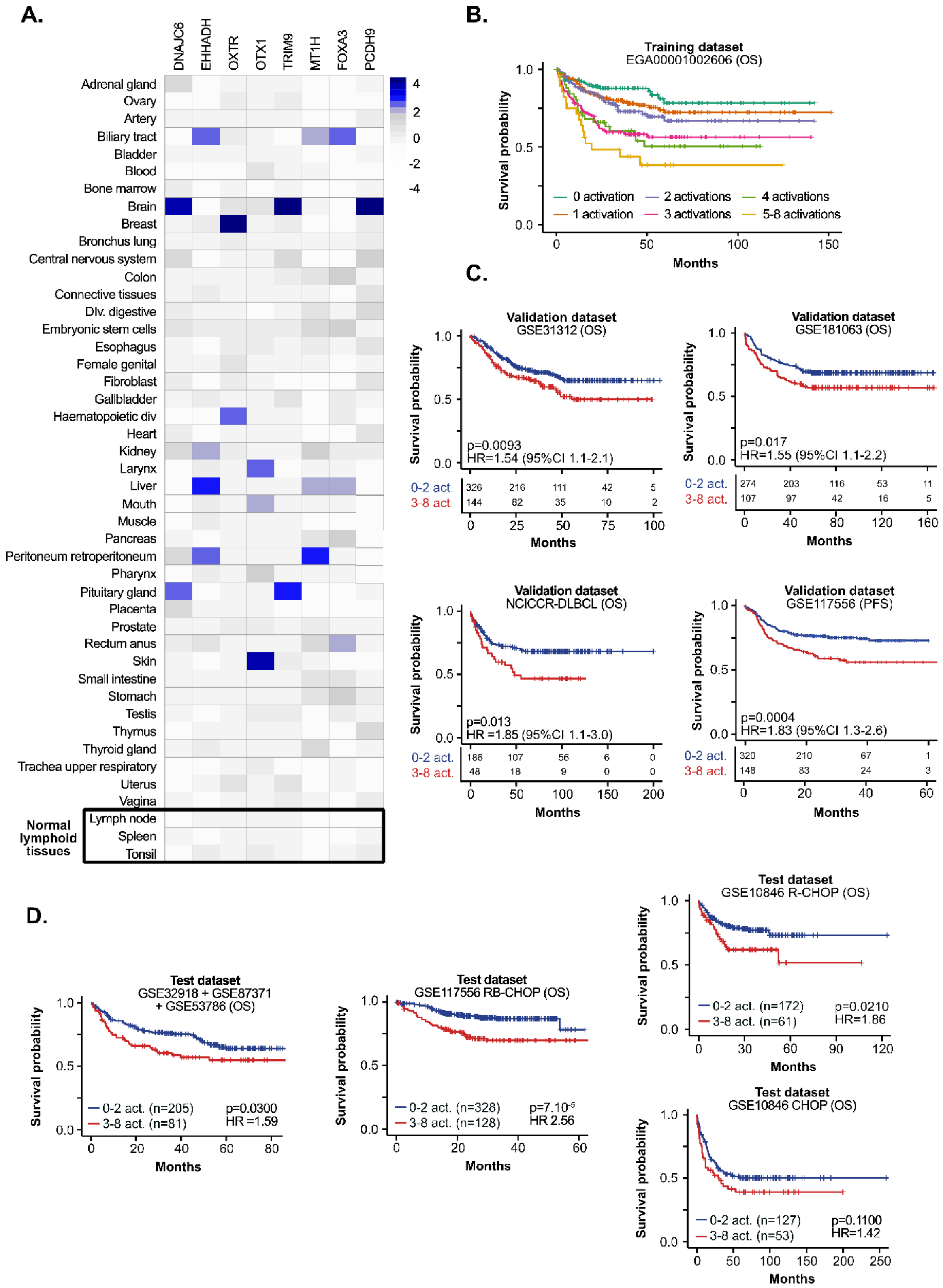


**Supplemental Figure 1. LymphoGEC DLBCL prognosis signature identification and validation. A.** Heatmap representation of LymphoGEC gene expression profiles in normal tissues (extracted from TSG3 database) illustrating their tissue-specific expression profile. **B.** Kaplan-Meier overall survival (OS) curves comparing patients presenting cumulative (from 0 to 5-8) LymphoGEC gene activation in EGA00001002606 dataset, global Cox model p-value = 1.0e$^{-09}$. **C-D.** Kaplan-Meier overall survival (OS) and progression-free (PFS) curves comparing patients presenting 0-2 and 3-8 LymphoGEC gene activation in all other validation (C) and test (D) datasets. Cox model p-values and hazard ratios (HR) including 95% confidence interval (95%CI) are reported.

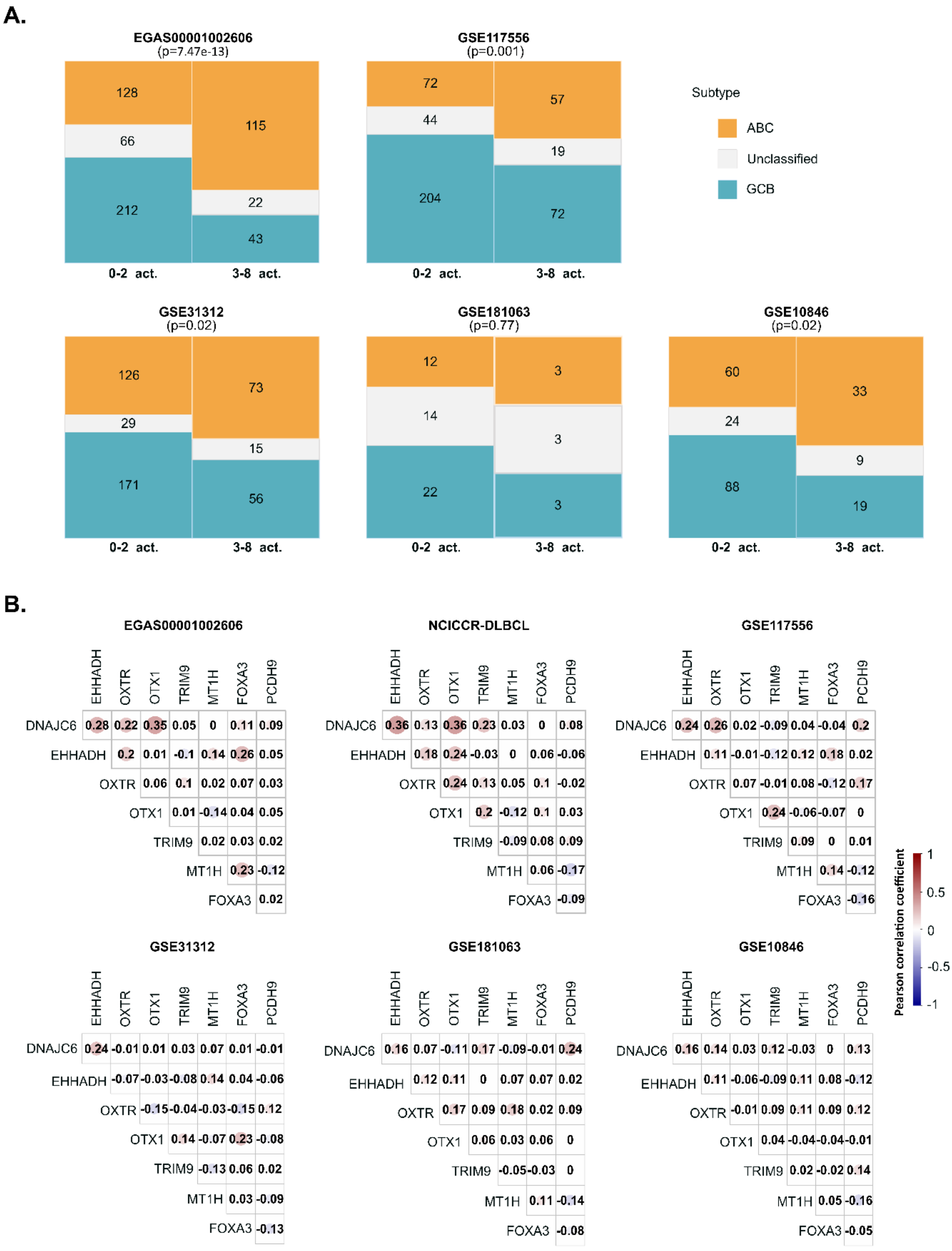


**Supplemental Figure 2. Correlation analyses of LymphoGEC expression profiles. A.** Mosaic plot showing the distribution of DLBCL molecular subtypes (ABC, GCB and unclassified) according to number of LymphoGEC reactivation (0-2, 3-8) in 5 of the DLBCL datasets as indicated. Chi square test's p-value are reported. **B.** Pearson correlation analysis of the 8 LymphoGEC genes in 6 of the DLBCL datasets as indicated. Pearson coefficients are reported.

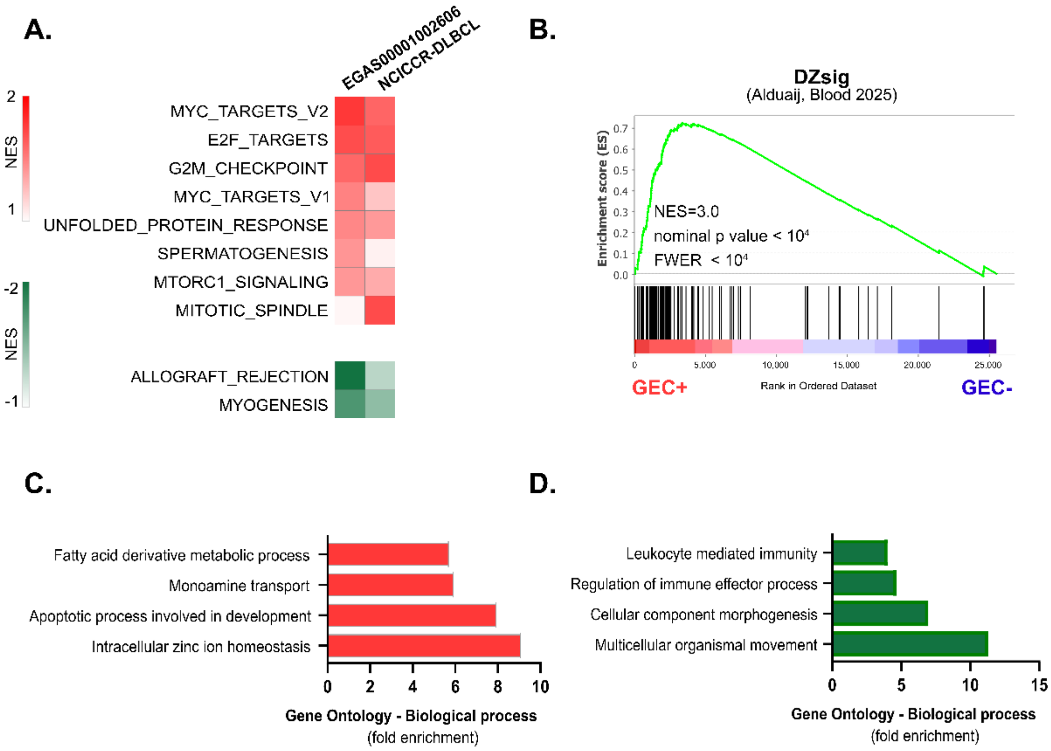


**Supplemental Figure 3. Impact of LymphoGEC reactivation on LBCL gene expression profiles**. **A.** Heatmaps representing normalized enrichment scores (NES) for GSEA signatures (hallmark database) with an FDR<0.05 in at least one of the EGA00001002606 and NCICCR LBCL datasets when comparing GEC+ (4-8 GEC activations) and GEC- (0 GEC activations) cases. Signatures enriched in GEC+ group are represented in red, signatures enriched in GEC- group are represented in green. **B.** GSEA plots showing enrichment for the dark-zone signature (DZsig) in GEC+ cases, NES: Normalized Enrichment Score, FWER: Family-Wise Error Rate. **C-D.** Fold enrichment for the top 4 Gene Ontology Biological process categories (WebGestalt analysis toolkit) for the commonly upregulated (red, C) and downregulated genes (green, D) between GEC+ and GEC- LBCL cases in the EGA00001002606 and NCICCR-DLBCL dataset.

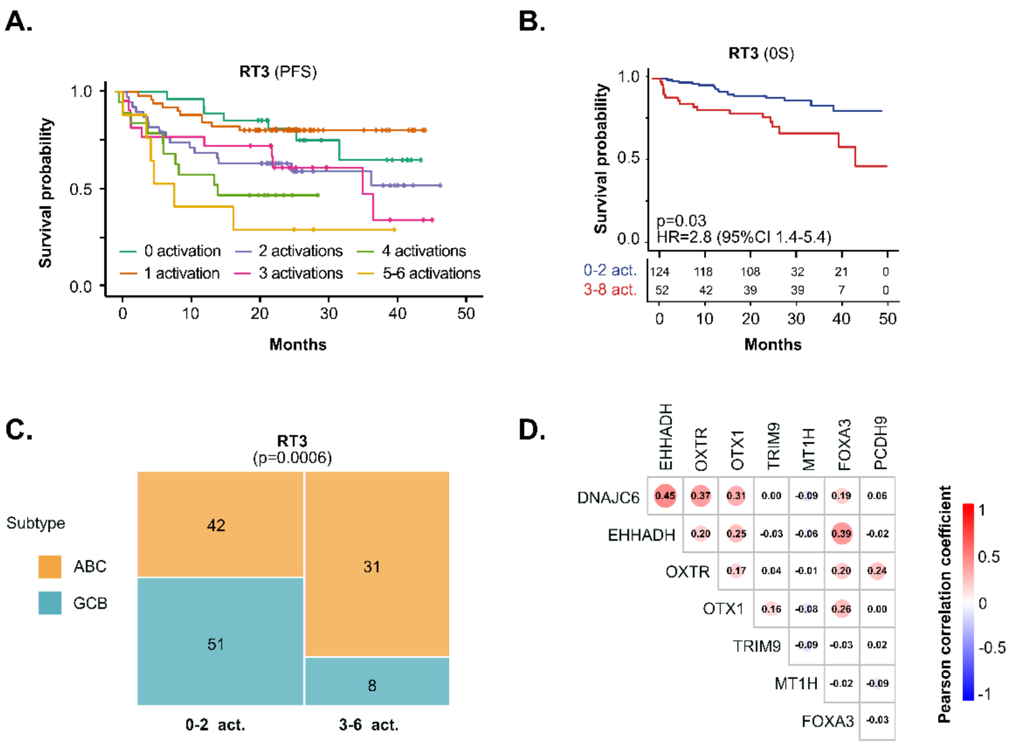


**Supplemental Figure 4. LymphoGEC validation in the RT3 cohort. A.** Kaplan-Meier progression free survival (PFS) curves comparing patients presenting cumulative (from 1 to 5-6) LymphoGEC gene activation in RT3 cohort, global Cox model p-value = 0.002. **B.** Kaplan-Meier overall survival (OS) curves and risk table comparing patients presenting 0-2 and 3-6 LymphoGEC gene activation in the RT3 cohort. Cox model p-values and hazard ratios (HR) including 95% confidence interval (95%CI) are reported, **C.** Mosaic plot showing the distribution of DLBCL molecular subtypes (ABC and GCB) according to number of LymphoGEC reactivation (0-2, 3-6) in the RT3 cohort. Chi square test's p-value is reported. **D.** Pearson correlation analysis of the 8 LymphoGEC genes in the RT3 cohort. Pearson coefficients are reported.
